# Hemophagocytic lymphohistiocytosis (HLH) in 2025 Dengue outbreak in Chittagong, Bangladesh

**DOI:** 10.64898/2026.02.14.26346308

**Authors:** Mohammed Nasir Uddin, Sk Md Faisal Abdullah, Nayan Dhar, Nahian Khan, Rajat Sanker Roy Biswas

## Abstract

**Introduction:** Hemophagocytic lymphohistiocytosis (HLH) is a serious condition induced by Dengue virus which becomes fatal if not detected early and treated appropriately. So objectives of the present study are to observe the different patterns of presentations, clinical features and outcome of HLH induced by Dengue.

**Methods:** In this observational study, 14 patients admitted and diagnosed HLH as per diagnostic criteria, were included after informed written consent. Study conducted in a period of six months from 01/07/2025 to 31/12/2025. All patients were followed up till discharge. After collection, all data were analyzed by Microsoft Excel 2010. Ethical clearance was taken from Ethical Review Board of the Medical College.

**Results:** Among 14 cases, male were more affected then the female (78.6% VS 21.4%) and majority were in between 20 to 50 years age groups. Clinical data showed, all 14 cases had fever for >7 days, joint pain 3(21.4%), headache 11(78.6%), skin rashes 10(71.4%), retro-orbital pain 2(14.3%), vomiting 11(78.6%),bleeding 10(71.4%), cough 4(28.6%), loose motion 9(64.3%), abdominal pain 7(50.0%), anorexia 2(14.3%), Melaena 2(14.3%), jaundice 4(28.6%) and spleenomegaly 9(64.3%). One(7.1%) case had history of Hypertension. Laboratory data showed different level of Bi or Pancytopenia, high ferritin, high TG, low fibrinogen, raised liver enzymes and low sodium. Dengue RT PCR and serology results showed 8(42.9%) cases were both IG M and Ig G dengue antibody positive, 6 cases were RT PCR positive, 2 cases were IgM and another 4 cases were IgG positive. Outcome of patients revealed, among all 14 cases12(85.8%) patients improved uneventfully and 2 were shifted to ICU where one improved and one died.

**Conclusion:** Dengue is prevailing for long time and different complications are evolving and HLH is a relatively newer incident among the dengue patients. Infection by different serotypes at different time or multiple dengue serotype infection may be related with HLH and it might be a future subject to explore and to evaluate.

## Introduction

Hemophagocytic lymphohistiocytosis (HLH) is an uncommon fatal disease of otherwise normal but hyperactive lymphocytes and histocytes [1]. HLH could be primary (hereditary) or secondary (acquired). Fever, hepatosplenomegaly, lymphadenopathy, and neurologic dysfunction are among the common symptoms of HLH. The diagnosis of HLH is based on clinical and biochemical findings

HLH is primarily caused by mutations in genes that are responsible for the production of cytotoxic T cells and natural killer (NK) cells. These are used to kill cells infected with pathogens like the Epstein-Barr virus (EBV) or the dengue virus [2]. Secondarily, it can be triggered by malignant conditions like acute lymphocytic leukemia, acute myeloid leukemia, Bcell lymphoma, T-cell lymphoma, and myelodysplastic syndrome or connective tissue diseaseslike juvenile idiopathic arthritis, juvenile Kawasaki disease, systemic lupus erythematosus (SLE), Still’sdisease - juvenile and adult-onset, and rheumatoid arthritis (RA), which have a tendency to damage the immune system. [3]

Infection caused by EBV, Human Immune-deficiency virus (HIV), Dengue virus, Cytomegalovirus (CMV), bacteria, fungi, protozoa, and now possibly severe acute respiratory syndrome Coronavirus 2 (SARS-COV-2) can be a cause of secondary HLH and there are some iatrogenic causes like organ transplantation, chemotherapy, or immunosuppressive therapy can also result in secondary HLH[4,5]

Diagnosis of HLH can be established either on the confirmation of the molecular diagnosis, e.g., pathological mutations of PRF1, UNC13D, Munc18-2, Rab27a, STX11, SH2D1A, or BIRC4, or on five or more of the following clinical findings: fever ≥ 38.5°C, splenomegaly, cytopenia, hypertriglyceridemia, hemophagocytosis in bone marrow, spleen, lymph nodes, or liver, low or absent NK cell activity, elevated CD25 (α-chain of SIL-2 receptor), and ferritin> 500 μg/L.[6]

Although HLH is now being increasingly detected in clinical practice due to improved understanding on the part of physicians, pathologists and microbiologists; still much work remains to raise awareness, explore treatment options and improve outcome of this complex condition. We want to observe report of dengue associated HLH who had a successful outcome following timely diagnosis and appropriate intervention..

## Methods

This is a prospective observational study, conducted in the Department of Medicine at CMOSHMC during a period of six months from 01/07/2025 to 31/12/2025. Patients admitted with a diagnosed case of Dengue fever either serologically or by molecular process with fever lasting more than 7 days were observed and investigated. After initial evaluation patients were observed for loose motion, rash, joint pain, headache, retro orbital pain, conjunctivitis, vomiting, bleeding, cough, abdominal pain or anorexia. Complete Blood Count, S. Ferritin, Triglyceride, Fibrinogen, ALT, AST and Serum Sodium level were advised for evaluation. Genetic and molecular diagnosis and bone marrow examinations could not be done due to lack of expertise and infrastructure. Patients were diagnosed as HLH as per criteria of HLH 2004 were included in the study except Genetic and molecular diagnosis and bone marrow examinations. According to this criteria, diagnosis of HLH can be established either on the confirmation of the molecular diagnosis, e.g., pathological mutations of PRF1, UNC13D, Munc18-2, Rab27a, STX11, SH2D1A, or BIRC4, or on five or more of the following clinical findings: fever ≥ 38.5°C, splenomegaly, cytopenia, hypertriglyceridemia, hemophagocytosis in bone marrow, spleen, lymph nodes, or liver, low or absent NKcell activity, elevated CD25 (α-chain of SIL-2 receptor), and ferritin> 500 μg/L, High TG or Fibrinogen level[6]. Patients unwilling to be included in the study or HLH unrelated to dengue were excluded. As Dengue with HLH is a rare condition, all patients fulfilling the inclusion criteria were included in the study during the six months study period and we got 14 cases of HLH related with dengue as per inclusion criteria. All data were collected in a case record form and after compilation,analysis was done by Microsoft Excel 2010. All patients eligible for the study were explained about the aims and objectives of the study and prior informed written consent was taken from all subjects. An ethical clearance from the ERB, CMOSHMC[CMOSHMC/IRB/2025/11] was obtained before starting of the study.

## Results

Table 1 showing male were more affected then the female 78.6% VS 21.4%) all age group from under 20 to more than 50 years patients were the victims and those had various occupations.

**Table 1:** Demographic data.

|  |  | Frequency | Percent |
| --- | --- | --- | --- |
| Sex | Female | 3 | 21.4 |
|  | Male | 11 | 78.6 |
| Age group | <20 years | 2 | 14.3 |
|  | 21-30 years | 4 | 28.6 |
|  | 31-40 years | 4 | 28.6 |
|  | 41-50 years | 2 | 14.3 |
|  | >51 years | 2 | 14.3 |
| Occupation | Business | 1 | 7.1 |
|  | House wife | 1 | 7.1 |
|  | Housewife | 1 | 7.1 |
|  | Retired | 1 | 7.1 |
|  | Service | 6 | 42.9 |
|  | Student | 4 | 28.6 |

Table 2 showing different clinical data where rash was 10(71.4%), all 14 patients had fever for >7 days, joint pain 3(21.4%), headache 11(78.6%), retroorbital pain 2(14.3%), vomiting 11(78.6%), Bleeding 10(71.4%), cough 4(28.6%), loose motion 9(64.3%), abdominal pain 7(50.0%), anorexia 2(14.3%) H/O HTN 1 7.1%, Melaena 2(14.3%) splenomegaly 9(64.3%) and jaundice 4(28.6%).

**Table 2:** Clinical data.

| Variables | Frequency | Percent |
| --- | --- | --- |
| Rash | 10 | 71.4 |
| Fever >7 days | 14 | 100 |
| Joint pain | 3 | 21.4 |
| Headache | 11 | 78.6 |
| Retro-orbital pain | 2 | 14.3 |
| Vomiting | 11 | 78.6 |
| Bleeding | 10 | 71.4 |
| Cough | 4 | 28.6 |
| Loose motion | 9 | 64.3 |
| Abdominal pain | 7 | 50.0 |
| Anorexia | 2 | 14.3 |
| H/O HTN | 1 | 7.1 |
| Melaena | 2 | 14.3 |
| Splenomegaly | 9 | 64.3 |
| Jaundice | 4 | 28.6 |

Table 3 showing laboratory data where different levels of cytopenias, higher ferritin, TG, fibrinogen, raised liver enzymes and low sodium were found

**Table 3:** Laboratory findings.

| Variables | N | Minimum | Maximum | Mean | Std. Deviation |
| --- | --- | --- | --- | --- | --- |
| S. Ferritin | 14 | 285.30 | 15050.00 | 5454.8221 | 5030.69958 |
| Triglyceride | 14 | 200.00 | 568.00 | 351.8571 | 122.56292 |
| Fibrinogen | 14 | .44 | 3.21 | 1.1600 | .83274 |
| ALT | 14 | 33 | 1544 | 382.64 | 449.461 |
| AST | 14 | 108 | 1789 | 576.36 | 571.955 |
| Serum Sodium | 14 | 123 | 143 | 134.14 | 4.897 |
| Total count | 14 | 2400 | 10900 | 5757.14 | 2770.924 |
| PLT count | 14 | 9000 | 200000 | 67285.71 | 69913.604 |
| Hemoglobin | 14 | 8.60 | 15.90 | 12.8000 | 1.58308 |

Table 4: Dengue RT PCR and serology results showed 6(42.9%) cases were both IG M and Ig G dengue antibody positive, 2 cases were RT PCR positive, 4 cases were IgM and another 4 cases were IgG positive.

**Table 4:** Dengue serology.

| Test name | Frequency | Percent |
| --- | --- | --- |
| Ig M for Dengue positive | 2 | 14.3 |
| Ig G for Dengue Positive | 4 | 28.6 |
| Both Ig M and Ig G positive | 8 | 57.72 |
| RT PCR for Dengue positive | 6 | 42.9 |

Table 5 depicts outcome of study patients where 12(85.8%) patients improved uneventfully and 2 cases were shifted to ICU where one improved and one died.

**Table 5:** Outcome of the HLH patients.

| Outcome | Number | Percent |
| --- | --- | --- |
| Improved uneventfully | 12 | 85.8 |
| Sifted to ICU | 2 | 14.3 |
| Improved and discharged | 1 | 7.15 |
| Death | 1 | 7.15 |

## Discussion

Accurate and early diagnosis of the prevalence and distribution of HLH within the population are difficult to obtain due to various factors, the most apparent of which is imprecise diagnostic criteria and the presence of multiple confounding medical illnesses at the time of diagnosis. The most comprehensive data regarding primary HLH comes from a Swedish national registry that collected data from 1987 to 2006 and demonstrated a yearly incidence of roughly 1.5 per million.[7]

Primary HLH presents in early childhood due to genetic mutations impairing the interaction between NK cells, CD8+ cytotoxic T-cells, and antigen-presenting cells. But we cannot diagnose it confidently due to lack of diagnosis facilities and expertise. Again Secondary HLH presents in adults with a mean age of 50 in response to an acute illness trigger rather than an underlying genetic mutation involve secondary HLH include infections (eg, tuberculosis, fungal, and histoplasma both with and without HIV) and malignancy.

Infection caused by EBV, human immune deficiency virus (HIV), dengue virus, cytomegalovirus (CMV), bacteria, fungi, protozoa, and now possibly severe acute respiratory syndrome coronavirus 2 (SARS-COV-2) can be a cause of secondary HLH[8].

In the present study all patients who were diagnosed as dengue but suffering from fever for a long duration with more than five criteria of HLH diagnosed fulfilled. It is was seen that a bigger number of patients had both Ig M and Ig G positivity for dengue. These might payed an immunological pathogenesis which might be explored in future.

Whether is it is an inherited HLH or an acquired one, it shares common pathophysiology. In a simplified way, it is suggested that it leads to an unchecked immune response while encountering the triggers. The hallmark of HLH is NK cell cytotoxicity. Familial HLH is linked to granule-dependent cytotoxicity. The inability to eliminate antigen-presenting cells and infected ones leads to uncontrolled proliferation and activation of the immune response with abundant cytokines. The clinical picture of HLH is due to the invasion of these cells in the organs and releasing more cytokines. Interleukin 1 (IL-1), IL-6, and tumor necrosis factor (TNF)-alpha cause the fever. The suppression of hematopoiesis by TNF-alpha and TNF-gamma results in cytopenia. Activated macrophages release ferritin and plasminogen activator, which leads to hyperfibrinolysis.[9]

In the study regarding different clinical data where rash was 10(71.4%), all 14 patients had fever for >7 days, joint pain 3(21.4%), headache 11(78.6%), retro-orbital pain 2(14.3%), vomiting 11(78.6%), orificial bleeding 10(71.4%), cough 4(28.6%), loose motion 9(64.3%), abdominal pain 7(50.0%), anorexia 2(14.3%) H/O HTN 1 7.1%, Melaena 2(14.3%) spleenomegaly 9(64.3%). Again laboratory data laboratory data showed different levels of cytopenias, higher ferritin, TG, low fibrinogen, raised liver enzymes and low sodium were found. These finding were consistent with a previous study. In a study done among 122 cases, fever, spleenomegaly, hepatomegaly, anemia, thrombocytopenia, and serum ferritin ≥500 μg/L were likely to report by articles representing by large sample size. The pooled proportion of these findings were: fever (n = 118) (97.2% [95% CI: 91.6□99.1], hepatomegaly (n = 108) (70.2% [95% CI: 51.1□84.2]), spleenomegaly (n = 110) (78.4% [95% CI: 54.5□ 91.6]), thrombocytopenia (n = 115) (90.1% [95% CI: 63.7□97.9]), anemia (n = 113) (76.0% [95% CI: 56.4□88.5]), serum ferritin ≥500 μg/□ (n = 102) (97.1% [95% CI: 90.5□99.2]). Prolonged thrombocytopenia (thrombocytopenia for 10 days or more) was found in 55.6% of 45 patients whose data was available [95% CI: 40.96□69.2]. Coagulopathy was found in 91.2% of 43 patients.[10]

In the present study all diagnosed HLH patients were treated with Dexamethasone which is considered very beneficial. 13 out of 14 patients with severe disease survived after the administration of dexamethasone in our study. Same results were found in a study done earlier[11] Literature is suggestive that the use of steroids in the HLH in severe dengue patients requires more study.[11,12] It is observed that the levels of ALT, AST, creatinine, LDH, and ferritin remain significantly higher in severely affected patients. Increasing AST and peak ferritin levels are linked with higher fatality. Physicians must consider HLH in dengue-infected patients if they observe persistent fever, abnormal mental state, cytopenia with organ issue, and, importantly, ferritin greater than 10,000 μg/L.[13]

In conclusion we can say that though it is a study of small sample but HLH might be a future threat for the community as all dengue serotypes are prevailing in the country. Heterotypic Dengue infection in the same year or in the subsequent year may increase the burden of the disease and may increase the fatality. Multicenter largescale study is needed to get the more precise data which might help to get good outcome.

## Data Availability

Data are available on valid request

